# Assessment of effectiveness of a COVID-adapted diagnostic pathway for colorectal cancer to mitigate the adverse impact on investigation and referrals

**DOI:** 10.1101/2020.11.23.20236778

**Authors:** Janice Miller, Yasuko Maeda, Stephanie Au, Frances Gunn, Lorna Porteous, Rebecca Pattenden, Peter MacLean, Colin L Noble, Stephen Glancy, Malcolm G Dunlop, Farhat V N Din

## Abstract

**Objectives:** The Coronavirus-19 (COVID-19) pandemic continues to impose formidable challenges on healthcare services. The dramatic curtailment of endoscopy and CT colonography capacity has adversely impacted on timely diagnosis of colorectal cancer (CRC). We describe a COVID-adapted pathway rapidly implemented to mitigate risk and maximise cancer diagnosis in patients referred with symptoms of suspected CRC during the pandemic.

**Design:** The “COVID-adapted pathway” integrated multiple quantitative faecal immunochemical tests (qFIT), to enrich for significant colorectal disease. CT with oral contrast was used to detect gross pathology. Patients reporting ‘high-risk’ symptoms were triaged to qFIT+CT and the remainder underwent initial qFIT. Prospective data collection comprised referral category, symptoms, blood results, medical history, time to first test, qFIT and CT results.

**Setting:** Tertiary colorectal unit which manages over 500 cancer patients annually.

**Participants:** All patients referred as ‘urgent suspicious of cancer’ (USOC) were included. Overall 422 patients (median age 64 years, 220 females) were triaged using this pathway.

**Main outcome measures:** Outcomes comprised cancer detection frequency.

**Results:** Compared to the same time period (1^st^ April – 31^st^ May) in 2017-2019, we observed a 43% reduction in primary care referrals with suspected CRC (1071 referrals expected reducing to 609). Overall 422 patients (median age 64 years, 220 females) were triaged using this pathway. Most (84·6%) were referred as USOC. Of the 422 patients, 202 (47·9%) were triaged to CT and qFIT, 211 (50·0%) to qFIT only, eight (1·9%) to outpatient clinic, and one to colonoscopy. Fifteen (3·6%) declined investigation and seven (1·7%) were deemed unfit. We detected 13 cancers (3·1%); similar to the mean cancer detection rate from all referrals in 2017-2019 (3·3%).

**Conclusions:** The response to the COVID-19 pandemic resulted in a marked reduction in referrals and cessation of key diagnostic services. Although this COVID-adapted pathway mitigated the adverse effects on diagnostic capacity, the overall reduction in expected diagnoses is very substantial. It is clear that the adverse impact of measures taken to constrain the pandemic will lead to many undetected cancers due to the decrease in referrals.

**Trial registration:** Not applicable

## Introduction

The collateral damage from the COVID-19 pandemic will have a lasting impact on colorectal cancer (CRC) survival^1,2^. The aerosol generating potential of endoscopy, coupled with COVID-19 faecal presence^3–6^, led to multiple colorectal and gastroenterology societies suggesting immediate cessation of all but emergency colonoscopy^7,8^. This resulted in suspension of the UK national bowel screening programme, which detected 20·3-20·9% of all colorectal cancers (CRC) since the start of bowel cancer screening in our unit in 2018. A similar rationale led to a pause in full CT colonography (CTC) suggested by the British Society of Gastrointestinal and Abdominal Radiology^9^. It was clear, through a combination of delayed presentation to primary care, decreased referral rates and lack of diagnostics, there would be an inevitable delay in CRC diagnosis.

Given the foreseeable fallout from a tertiary unit managing over 500 colorectal cancer patients annually, we assembled a team to ensure rationing of available diagnostics was evidence-based and would further enrich the traditional symptom-based approach^10^. Whilst the sensitivity of unprepared CT scans for CRC compared to CTC is lower (75-80% vs 95%), guidance advised deferral of luminal investigation in patients with a negative standard CT scan^9,11^. The other tool available for CRC detection was the quantitative faecal immunochemical test (qFIT) used for screening and as a triage tool in ‘low risk’ populations^12,13^. qFIT does enrich for bowel pathology but cannot be used as a ‘rule-out’ for CRC given the test sensitivity^14^.

Hence, we designed and rapidly implemented a pragmatic approach to mitigate risk and maximise cancer diagnosis utilising plain CT and qFIT^15^. A collaborative approach involved colleagues from biochemistry, radiology, general practice (GP) and gastroenterology. GP referrals of ‘urgent suspicion of cancer’ patients (USOC) were triaged daily by colorectal consultants using age, symptoms (‘high-risk vs low-risk’) and haemoglobin to prioritise those most likely to have CRC^13^. Patients were triaged to the limited diagnostics accounting for the limited availability of both surgical and oncological services, staff and appropriate personal protective equipment (PPE). Here, we present the outcomes of this approach and assess the extent to which our COVID-adapted pathway for USOC patients mitigated the risk of delayed diagnosis due to the pandemic.

## Methods

To support referrals for USOC patients during the pandemic a triage process was developed based on local endoscopy and imaging capacity and qFIT testing. This was done to channel the type and timing of investigations, interspersed with safety-netting mechanisms including telephone or outpatient assessment and prioritisation to urgent colonoscopy as appropriate. A summary of our COVID-adapted pathway is shown in Figure 1^15^.

**Figure 1.**
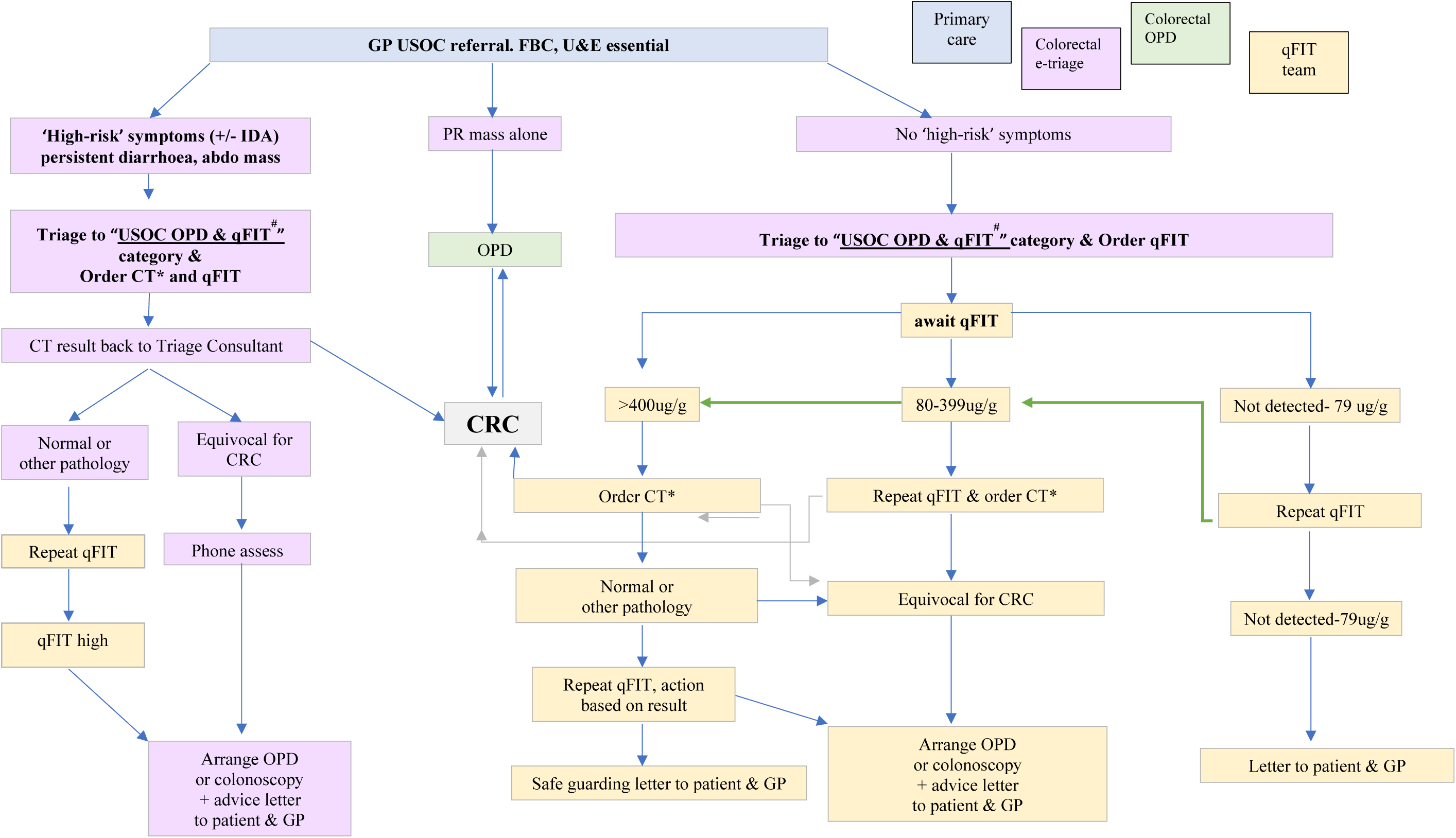
NHS Lothian COVID adapted colorectal cancer pathway. A summary of the COVID-adapted pathway. Patients were triaged by colorectal consultants with information provided from general practice. They proceeded through the pathway in a step-wise fashion being stratified by qFIT results. CT: computed tomography scan, CRC: colorectal cancer, IDA: iron deficiency anaemia, qFIT: quantitative faecal immunochemical test, OPD: outpatients department, USOC: urgent suspected of cancer.

### Eligibility criteria and index testing

Through direct communication GPs were advised to continue to refer USOC patients or those with symptoms suggestive of serious lower gastrointestinal (GI) disease.

All patients referred with USOC symptoms (palpable abdominal mass, persistent change in bowel habit to looser stool not just simple constipation, repeated rectal bleeding without an obvious benign anal cause or blood mixed in with the stool, abdominal pain with weight loss +/- iron deficiency anaemia [IDA]) consecutively entered the pathway between 1^st^ April to 31^st^ May 2020. All patient referrals were electronically triaged by colorectal consultants to one of three arms:

1. ‘High-risk’ symptoms +/- IDA: In this patient group a qFIT and CT minimal preparation scan were requested at the same time. Scans were reported by consultant radiologists as being ‘grossly normal’, ‘equivocal’ or ‘definite cancer’. All equivocal CT findings were double-reported by a second consultant radiologist. Those with an elevated qFIT but negative CT underwent repeat qFIT testing +/- colonoscopy.
2. Palpable rectal mass: These patients were seen in person at clinic. In the absence of a mass, qFIT tests were ordered to inform the next investigation.
3. Those with ‘lower risk’ symptoms underwent qFIT testing only initially. Patients were then stratified according to qFIT values to enrich for those most likely to have serious bowel pathology.

It would not have been pragmatic to use 10μg/g as the threshold for investigation given data from several health boards suggests the positivity rate is ∼23%. The initial threshold for further investigation was therefore based on the Scottish bowel screening guidelines (80μg/g)^16^. All patients who returned a test <80μg/g underwent repeat testing. Following two qFITs <80μg/g, and a negative CT for some patients, safety-netting advice was given. For all patients who returned tests at an intermediate level (80-399μg/g) a repeat qFIT test and CT was ordered. All those with a qFIT >400μg/g proceeded to CT without a second qFIT result. CT colon minimal preparation was a plain CT with the patient taking oral contrast at home and without rectal insufflation.

Colonoscopy was only performed on those patients with the most urgent need e.g. those with a CT result highly suspicious but not diagnostic for cancer that required a biopsy.

### Safety-netting and post-acute pandemic plans

Safety-netting protocols were enabled to allow for further clinical assessment if symptoms persisted or worsened. Patients in the ‘low-risk’ symptoms arm who had negative qFITs were reassured via letter their results suggested a low residual risk of cancer but further investigation based on symptoms may be required later. Patients were not discharged from the pathway on the basis of a single qFIT test alone. Investigations in vulnerable patients who met the referral criteria but were shielding were deferred based on patient preference after telephone consultation^17^. The pathway was adapted following the return of limited access to CTC and colonoscopy in June 2020. Due to the initial wide variability observed in double-testing all patients with two negative qFITs were offered a CT-minimal preparation scan. Those with tests of 10-399μg/g underwent CTC and those >400μg/g were referred for colonoscopy.

### Statistical analysis

All pathway patients were prospectively entered into a Microsoft EXCEL spreadsheet. Data was collected on entry route, triage category, demographics, presenting symptoms, blood results and past medical history. Results of all qFITs and colorectal investigations were recorded when complete. Given the limited access to CTC and colonoscopy there was no reference standard. Cancer detection rates during the pandemic were therefore compared to those from previous years where patients had undergone the standard diagnostic pathways. All clinical information was available to those reviewing both qFIT and CT results. All patients (including those who did not have a complete set of tests) were included in the analysis. Statistical analysis was performed using software R version 4.0 (http://www.R-project.org) with appropriate packages. Shapiro-Wilk’s method was used to test for normality. Non-parametric data are presented as median and interquartile range (IQR). Variability in two qFIT tests was measured using Pearson’s correlation coefficient.

### Patient and public involvement

Patients and the public were not involved in the design or implementation of this pathway.

## Results

### Patient demographics and initial triage outcomes

The first iteration of the COVID-adapted pathway ran from 1^st^ April to 31^st^ May 2020. There were 422 patients included, median age 64 years (55-74) with 220 being female. Patients were predominantly referred under the USOC category (357), with 48 urgent and 17 routine referrals being upgraded to USOC by the triaging colorectal consultant.

The time to first test was a median of 14 days (IQR 10-18) and there was no difference between testing time in CT (median 15 days, IQR:10-24) or qFIT (median 13 days, IQR:10-17). The median time from referral to qFIT completion was 22 days (IQR: 15-37).

Of the 422 patients, 202 (47·9%) were triaged to CT and qFIT based on ‘high-risk’ symptoms, 211 (50·0%) to qFIT only based on ‘low-risk’ symptoms, eight (1·9%) straight to the outpatient clinic as ‘palpable mass PR’, and one to colonoscopy and qFIT due to the patient having undergone a recent CT with a suspicious finding requiring luminal investigation. Of the patients who entered the pathway 15 (3·6%) declined any investigation and seven (1·7%) were deemed unfit due to shielding, severe cognitive impairment or being admitted to hospital for other reasons at the time of testing. There were 82 (19·4%) patients who had an incomplete set of results; they did not return either qFIT test within six weeks despite pro-active encouragement from nursing teams or declined to attend for CT (17). Thirty-six of these patients were from the ‘high-risk’ arm. The COVID-adapted pathway detected 13 CRCs (3·1%). This was on par with our performance of a yearly 3·3% cancer detection rate from all referrals in 2017-2019. The flow of patients leading to cancer diagnosis is shown in Figure 2.

**Figure 2.**
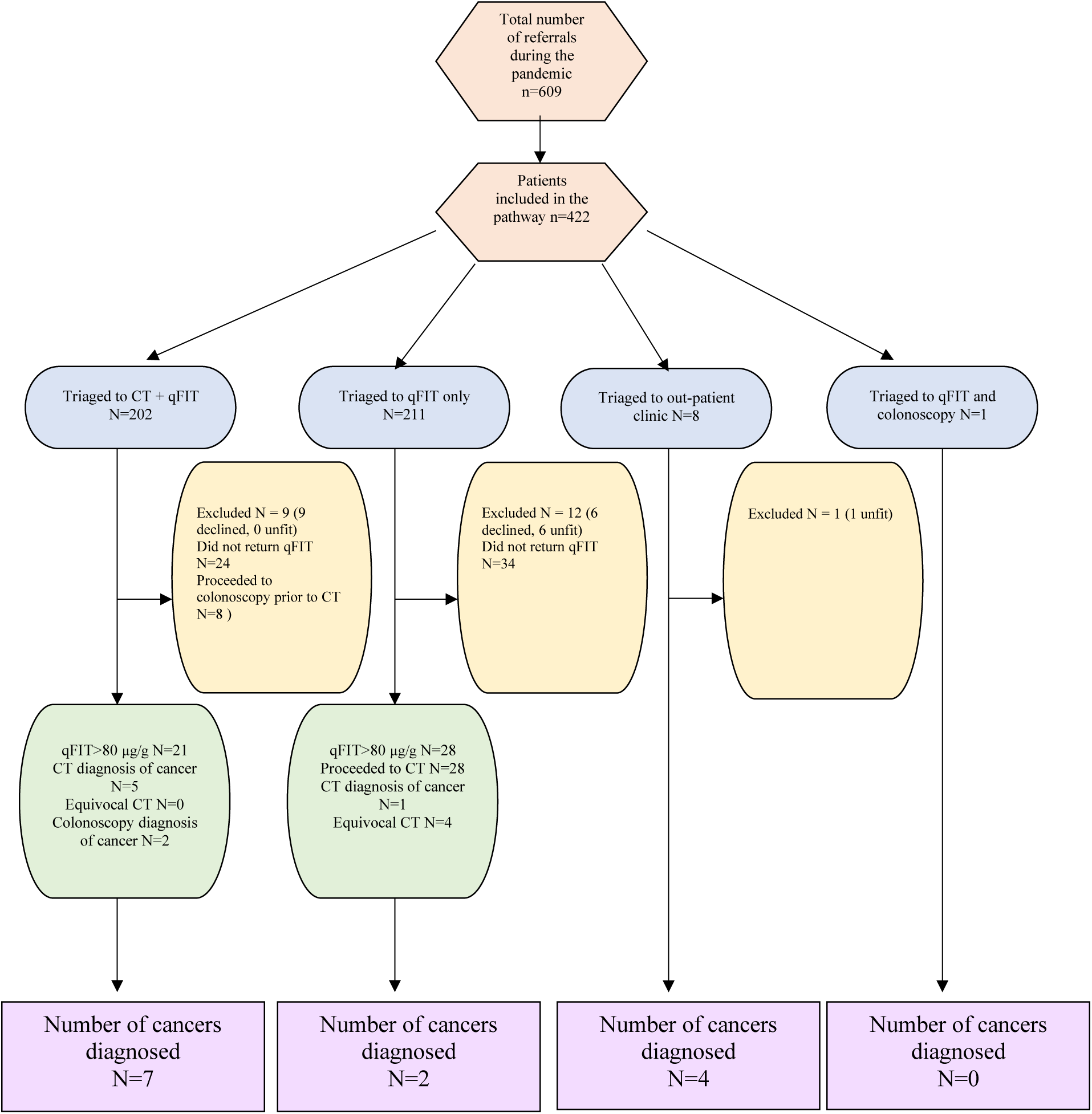
Flow of patients through the pathway leading to cancer diagnosis. Patients were diagnosed through a variety of routes, the maximal yield coming from those who had both initial CT and qFIT testing. With 50% being diagnosed from the outpatient clinic, the initial referral examination was deemed to be of great importance.

### Change in activity during the pandemic

The number of overall referrals and activities were compared with the previous three years (2017-2019) during the same period (April and May). The total number of referrals decreased by 43% from an average of 1071 to 609 during the pandemic, with a 79% reduction in urgent (324 to 69) and 64% reduction in routine (581 to 211) referrals. However, the number of USOC referrals increased by 40% (235 to 329). The decrease in overall referral numbers highlights that there may be as many as 50% more CRCs in the community that are yet to present or be referred. The average number of monthly cancer diagnosis in our unit was 44 during the last three years (2017-2019) whilst the average since February of this year was 30 per month. A comparison of referral rates from primary care over the four years is shown in Figure 3. A further 16 cancers were diagnosed as emergencies during April and May; an increase of 33% on 2019. Following the initial peak of the pandemic, referral numbers for June and July 2020 did not increase (377 and 337 respectively, down from 425 and 500 the previous year).

**Figure 3.**
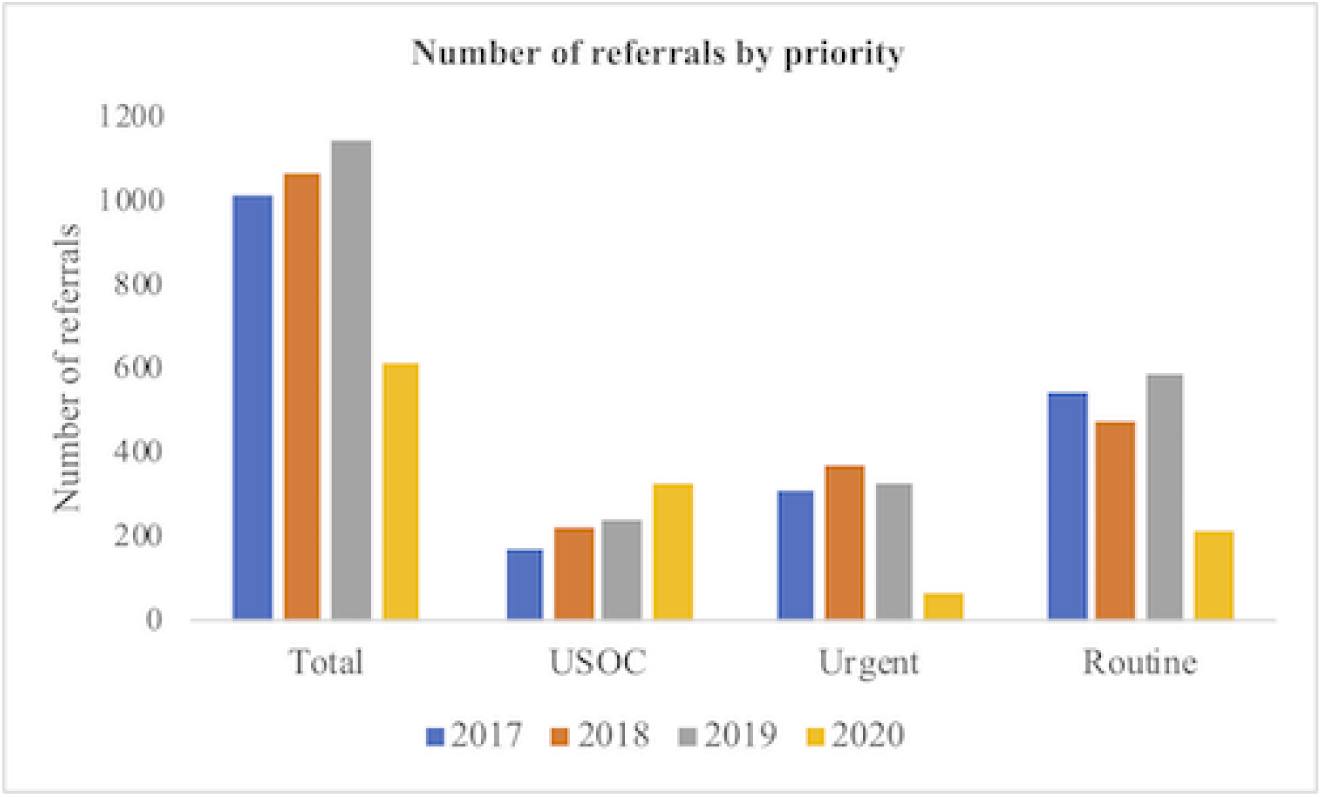
Number of referrals by priority 2017 - 2019. There was a marked decrease in the total number of referrals during the pandemic, with an increase in the number of USOC referrals.

### qFIT results

Of 422 patients, 366 (86·7%) completed at least one qFIT test. The majority of patients (266, 72.7%) had ‘not detected’ (<10ug/g) results, 51 patients (13·9%) had levels between 10-79µg/g, 18 patients (4·9%) had levels between 80-399µg/g and 31 patients (8·5%) had levels ≥400µg/g. The overall positivity rate was higher (27·3%) than our qFIT levels in symptomatic populations pre-COVID (22% from audit data) suggesting an already enriched population presenting and completing the tests.

263 patients (62·3%) completed at least one qFIT test and either a CT or colonoscopy as a definitive colorectal investigation on the pathway: 72·0% had an undetected qFIT, 12·6% had a result between 10-79µg/g, 6·3% had a result between 80-399µg/g and 9·1% had a qFIT at or over 400µg/g. Interestingly, we noted differences in the distribution of qFIT values between ‘high-risk’ and ‘low-risk’ groups. Contrary to intuitive assumptions, there were more patients with undetected qFITs in the ‘high-risk’ group (78·4%) compared to the ‘low-risk’ group (68·8%), whilst there were more patients with a qFIT over 400µg/g in the ‘low-risk’ group (9·5%) compared to the ‘high-risk’ group (7·0%). However, these differences were not statistically significant (p=0·19).

### Double qFIT testing

There is limited published data on the use of double qFIT testing to enrich or safety net CRC patients. Although multiple tests may enrich a few patients, we found considerable variation in interval double-test qFIT values (two tests at least two weeks apart). Two-hundred and twenty patients (52·1%) completed two qFIT tests of whom 184 (83·6%) had both qFIT values under 80µg/g. Eighteen patients (8·2%) had at least one qFIT >80µg/g which triggered a CT and a further 18 patients (8·2%) had both qFIT values >80µg/g. There were three cancers amongst those who had two <10µg/g qFIT tests. Two patients had both qFITs under 80µg/g and one patient had both qFITs above 80µg/g as shown in Figure 5. Pearson’s correlation coefficient was 0·63 showing moderate test-retest reproducibility, and the proportion of patients who had one qFIT <80µg/g and another qFIT >80µg/g was 8% (potential incremental diagnostic yield).

**Figure 4.**
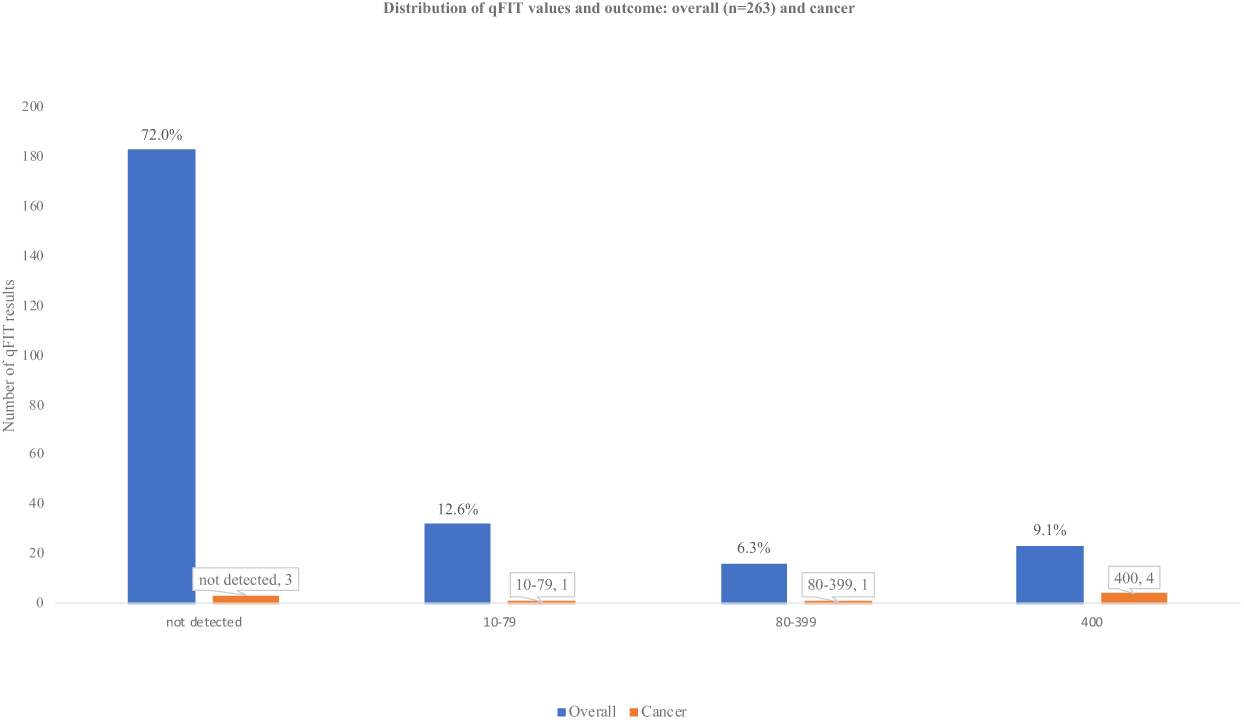
Distribution of qFIT results and overall outcome. The majority of patients had an undetected qFIT result. Despite this 3 cancers were diagnosed within this group.

**Figure 5.**
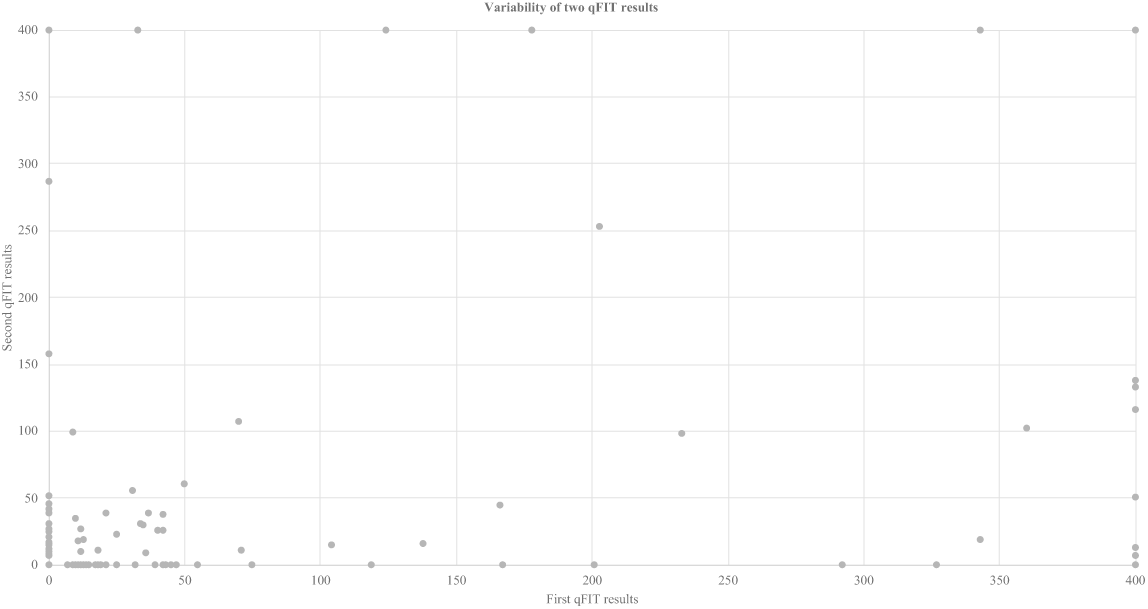
Distribution of double qFIT results. Double qFIT testing showed variability of results. Eighty-four percent of patients had both results <80 μg/g, 8% had one result <80 μg/g and one >80 μg/g and a further 8% had two results >80 μg/g. There were two cancers diagnosed in those with two qFIT’s <10 μg/g and one in a patient with two qFIT’s >400 μg/g.

### CT scan results

Of 265 CTs done, 241 were reported as normal, 15 as equivocal and nine as cancer: one of the 15 equivocal scans was found to be cancer and one of the patients with a scan reporting cancer was subsequently found to have benign disease. The rest of the ‘equivocal’ findings were investigated by endoscopy or CTC: five patients had diverticular disease, four patients were found to have polyps, two patients had colitis (UC, collagenous colitis), one was normal, one was presumed to be normal change due to previous surgery but awaiting MRI for further clarification, and one did not attend the endoscopy appointment.

### Cancer diagnosis

Thirteen CRCs were diagnosed overall. The median age at cancer diagnosis was 71 years (67-78) with six patients being female. The distribution of pathology and initial treatments are shown in Table 1 along with final TNM staging for those patients who have already proceeded to surgery. One patient had metastatic disease in the liver at presentation.

**Table 1.**
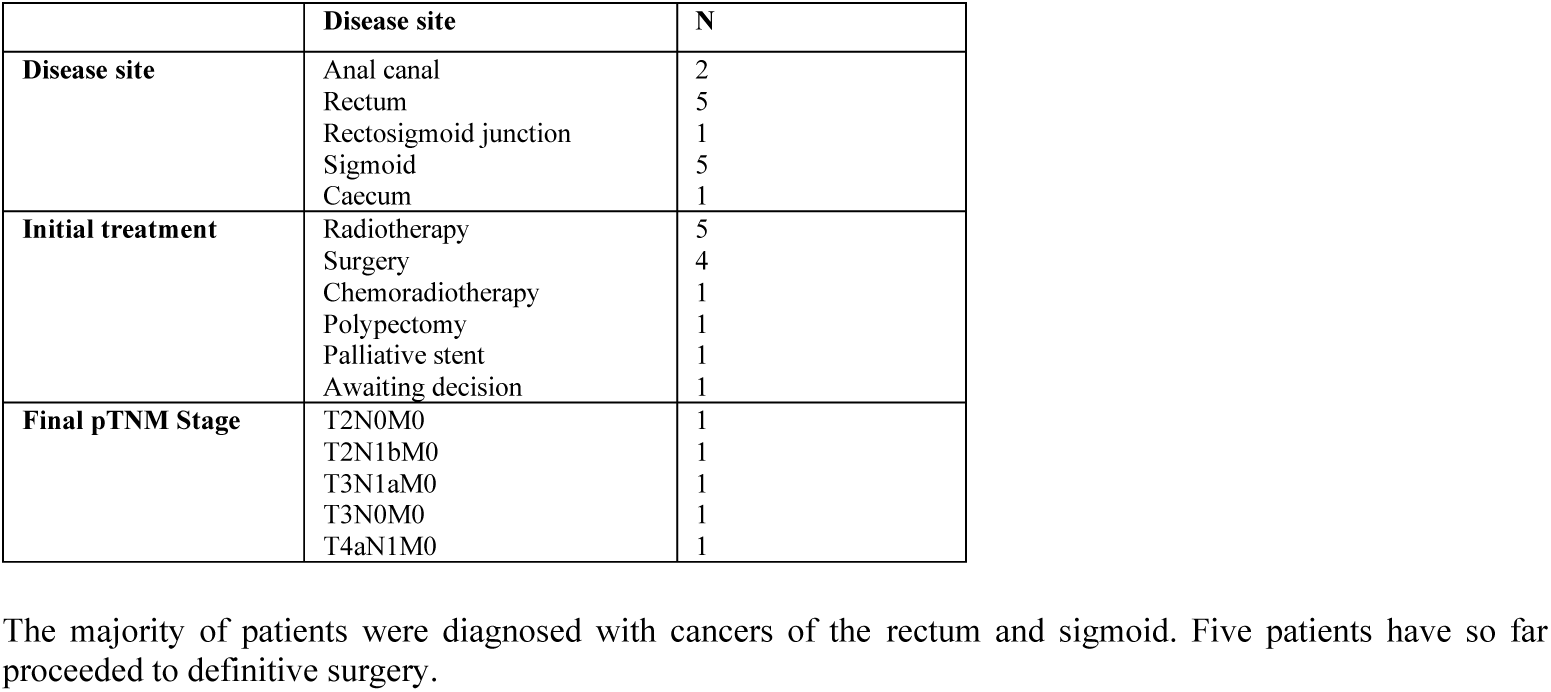
Pathological diagnoses in cancer patients.

|  | Disease site | N |
| --- | --- | --- |
| **Disease site** | Anal canal | 2 |
|  | Rectum | 5 |
|  | Rectosigmoid junction | 1 |
|  | Sigmoid | 5 |
|  | Caecum | 1 |
| **Initial treatment** | Radiotherapy | 5 |
|  | Surgery | 4 |
|  | Chemoradiotherapy | 1 |
|  | Polypectomy | 1 |
|  | Palliative stent | 1 |
|  | Awaiting decision | 1 |
| **Final pTNM Stage** | T2N0M0 | 1 |
|  | T2N1bM0 | 1 |
|  | T3N1aM0 | 1 |
|  | T3N0M0 | 1 |
|  | T4aN1M0 | 1 |

Seven cancers were identified from the CT and qFIT arm (3·5%), two cancers from the qFIT only arm (0·9%), and four cancers from the outpatient clinic arm (50%). These results highlight that our pathway design, based on symptoms, was appropriate given the greatest number of cancers came from the CT and qFIT arm (54%). Four cancers were diagnosed in the group with qFIT > 400µg/g, two with qFIT results of 10-79 µg/g and three patients with a qFIT result <10µg/g (Figure 4). Of those patients with a qFIT <10µg/g one patient had frequency and tenesmus, another loose stool, anaemia and weight loss and the final constipation and rectal bleeding. Given the smaller number of cancers diagnosed in this group there is potential that there are patients with undiagnosed cancer. Results of this will become apparent over time and once safety-netting investigations are complete. Two hundred and twenty-two (52·8%) out of 422 referrals had documentation of digital rectal examination by primary care, of whom 12 stated there was the possibility of a rectal mass. Eight of these patients were fast-tracked to the outpatient clinic. Of the remaining four, two were triaged to CT and qFIT and two to qFIT only. Only one patient with a palpable mass had a qFIT undertaken with the result being 21µg/g. Of the cancers that were anorectal a further two would have been diagnosed by rectal examination had it been done. There were a range of other diagnoses in those without the target condition that are summarised in Table 2.

**Table 2.** Alternate diagnoses from CT minimal preparation scans.

| Pathology | N |
| --- | --- |
| Ischaemic Colitis | 1 |
| Diverticulitis | 2 |
| Sigmoid polyps | 1 |
| Ulcerative Colitis | 1 |
| Recurrent breast cancer | 1 |
| Metastatic pancreatic cancer | 1 |
| Indeterminate lung lesion | 1 |
| Renal cyst | 1 |

A range of other diagnoses were also made as described above

### Adverse events from index tests

No adverse events were reported from the qFIT test. There were no perforations in those patients who proceeded to colonoscopy. One patient who was radiologically diagnosed as cancer was found to have complicated diverticular disease at final pathology but was symptomatic enough to require operation. One patient did not undergo CT scan due an iodine allergy. Two patients died following referral with neither starting the pathway. One patient had a CT-confirmed diagnosis of advanced cirrhosis and died with decompensated liver and cardiac failure 21 days following referral. Another died within nine days of referral of an unknown cause, they had a history of weight loss and anaemia but declined investigation for this the previous year.

## Discussion

We describe a novel COVID-adapted triage pathway for colorectal cancer detection which has mitigated risks for those referred with USOC symptoms during the pandemic. We have shown that it is possible to implement change rapidly if all stakeholders are committed and in doing so have highlighted the impact of delayed presentation to primary care on the potential number of missed cancers during this period. We have examined the impact of using qFIT and CT minimal preparation scanning on cancer detection rates and have demonstrated that the use of this novel COVID-adapted pathway has allowed us to match the expected cancer detection rate in those referred. The variability in double qFIT testing further emphasises the importance of clinical examination and adequate referral information. This pathway has helped to standardise treatment and optimise the balance between delivery of effective cancer care and minimisation of risks to patients and staff. We have developed a responsive framework which is ready to be utilised during further COVID-19 peaks.

These data have shown that there was a 43% reduction in referral numbers. This was possibly due to the blanket “lockdown” message by the government and media encouraging people to stay at home, compounded by the cessation of cancer screening services. Patient anxiety about attending hospital also increased in the elderly in particular – a group at high risk from COVID-19 but also most likely to have serious bowel pathology. It was expected that red flag symptoms such as rectal bleeding or a new lump would continue to present, however there was concern that more vague symptoms including change in bowel habit, symptoms of anaemia or weight loss would be dismissed by patients for fear of wasting doctors time for non-COVID-19 problems^18–20^.

The decrease in referral numbers was potentially increased by the reduced availability of face to face appointments in primary care and the huge shift towards telephone triage. As we have shown half of those patients referred with a rectal mass had a malignancy, suggesting many may have missed examination findings due to the increased use of remote consulting. This style of consultation is also less suited to those from lower socioeconomic backgrounds thus potentially increasing inequalities that are already exist in cancer care^21^.

It is expected that given the drop in referral numbers, there will be a significant number of patients in the community with CRC who are as yet undiagnosed. Large scale modelling studies have estimated >4700 deaths to be attributed to a three-month delay to all cancer surgery in England with further impacts on death and life years lost if diagnostic services are delayed in returning to normal^1,22^. There are likely to be many patients presenting with cancer symptoms or advanced cancer as emergencies in the future. For patients with localised CRC the risk of delay in presentation is not known, however, it may lead to upstaging of disease with one study predicting delays in diagnosis and management may lead to a 17·2% reduction in survival for those with stage one disease increasing to >29% in those presenting with stage three disease^2^. Delays in diagnosis not only lead to reduced survival but also potentially more morbid surgery and increased likelihood of neoadjuvant and adjuvant treatments. Patients presenting as emergencies are also more likely to require longer hospital stays and critical care^23^.

In June 2020, following the end of the initial pathway, referral numbers had not returned to the usual expected numbers. There was increased, but considerably constrained, access to endoscopy and CTC. Unless capacity is markedly increased there will continue to be delays to diagnosis and management of these patients which may persist over a long period of time. Our median time to index test was 14 days. Given the existing prolonged endoscopy waiting lists in many centres, the use of this pathway to triage patients will be required for the foreseeable future. COVID-19 has generated the opportunity to overhaul unwieldy triage systems and implement a stream-lined approach to patient assessment^24^. Hence, planning at the outset was undertaken to embed the pathway into routine service so as to reduce the never-ending burden on colonoscopy lists. There remains a residual risk for patients and safety-netting plans have been initiated to offer all patients who return a qFIT test <80 µg/g a second qFIT and CT-minimal preparation scan. There was considerable variation in initial triage category assigned by individual consultants and this should be standardised following the acute phase. qFIT is a useful test to prioritise access to endoscopy and CTC, with the caveat that symptomatic patients with a negative test will ultimately require investigation in the long term^25^.

### Limitations

It was not possible to validate the use of qFIT or CT minimal-preparation scan at this time given the inability to compare it to a reference standard. Our approach was based on the ability of the tests to diagnose gross pathology and as such this data will become available over time. We have focussed solely on the detection of overt CRC, with the detection rate of advanced polyps currently unknown. The sensitivity of qFIT for advanced adenomas has been shown to be low at 35·7%^26^. The value of qFIT is known to be significantly lower for more proximal adenomas and cancers compared to those found distally, with double qFIT testing enriching for pathology^26^. Although there were three cancers diagnosed in patients with a negative qFIT only one was a caecal malignancy, the others being sigmoid and anorectal lesions. The number of cancers diagnosed in those with two negative qFITs was too small to comment on whether double-testing enriched for pathology. Throughout the devolved nations different thresholds have been used to determine what constitutes a positive result for screening patients (80μg/g in Scotland, 120μg/g in England and 150μg/g in Wales)^27^. The threshold for determining an abnormal result is lower (10μg/g) in the symptomatic population^28^. It is used to enrich information and target patients with ‘low risk but not no risk’ symptoms^29^. Therefore, even a negative result may require further investigation in order to manage risk and demand. There are concerns therefore that the use of qFIT testing alone could miss cancers in some patients.

A small percentage of USOC referrals as shown lead to the eventual diagnosis of other cancers or pathologies. It is not known how a qFIT alone based triage system would impact on these diagnoses. We focussed on patients referred with USOC symptoms and did not include those referred via routine referrals or those under long-term polyp or genetic surveillance.

## Conclusion

The coronavirus pandemic led to a marked decrease in referrals and cessation of key diagnostic services. Public awareness campaigns should encourage those with ‘high-risk’ symptoms to come forward, stressing the importance of timely cancer diagnosis. Prioritisation of symptomatic patients through the use of qFIT and CT minimal preparation scans has been shown to be a rational approach to mitigate risk and prevent delay to treatment when access to endoscopy was limited. It is likely that the effect of the pandemic on non-COVID patients will out-weigh the current effects on health and economy.

## Data Availability

De-identified data is available from the corresponding author on request.

## Acknowledgments

We would like to thank colleagues who helped collating data and coordinating activities (Paulina Dluzynska and Nichole Jeffries) as well as colleagues in the departments of colorectal surgery, gastroenterology, radiology and biochemistry at the Western General Hospital, Edinburgh as well as colleagues in primary care for their help in triaging patients and timely reporting of results. MGD is funded by CRUK and MRC. FVND is funded by the Chief Scientist Office.

## Copyright

The Corresponding Author has the right to grant on behalf of all authors and does grant on behalf of all authors, an exclusive licence on a worldwide basis to the BMJ Publishing Group Ltd to permit this article (if accepted) to be published in BMJ editions and any other BMJPGL products and sublicences such use and exploit all subsidiary rights, as set out in our licence.

## Author contributions

JM, FG and YM helped run the pathway. JM, FG and SA collected data. JM,YM,MGD and FVND wrote the manuscript. YM, LP, RP, PM, CN, SG, MGD and FVND designed and set up the pathway. YM analysed data. Data was verified by JM, FVND and MGD. All authors critically appraised and approved the final manuscript.

## Ethical approval

Ethical approval to report pathway outcomes was not required given it was part of clinical care.

## Data sharing

Deidentified data will be available on request from the corresponding author.

## Transparency statement

The manuscript is an honest, accurate and transparent account of the study being reported.

## Summary box

### What is already known on this topic

The Coronavirus-19 (COVID19) pandemic has resulted in a severe limitation of access to endoscopy and CT colonoscopy services for patients with colorectal cancer due to fears surrounding its aerosol generating potential. Never before has there been the need to rapidly implement an alternative pathway for patients presenting with symptoms suspicious for colorectal cancer. Most data on the use of qFIT relates to the screening population with little known about its value in those who are symptomatic.

### What this study adds

We have described a novel COVID-adapted pathway to mitigate risk and instigate treatment for those presenting with symptoms of colorectal cancer during the pandemic. We combine the best available testing data (qFIT and CT minimal preparation scan) to provide a detailed assessment of the impact of reduced availability of diagnostic testing on cancer detection rates. Colorectal cancer is common. Reductions in referral numbers during the pandemic indicates the potential for missed diagnoses. The combination of qFIT and CT minimal preparation has the ability to mitigate risk and could be incorporated into global healthcare systems in the longer term in order to provide safety netting of patients until normal diagnostic services resume.

